# Harmonization of Multi-Center Diffusion Tensor Tractography in Neonates with Congenital Heart Disease: Optimizing Post-Processing and Application of ComBat

**DOI:** 10.1101/2021.10.01.21264443

**Authors:** Benjamin Meyers, Vincent K. Lee, Lauren Dennis, Julia Wallace, Vince Schmithorst, Jodie K. Votava-Smith, Vidya Rajagopalan, Elizabeth Herrup, Tracy Baust, Nhu N. Tran, Jill Hunter, Daniel J. Licht, J. William Gaynor, Dean B. Andropoulos, Ashok Panigrahy, Rafael Ceschin

**Affiliations:** Department of Radiology, University of Pittsburgh School of Medicine, Pittsburgh, PA; Division of Cardiology, Department of Pediatrics, Children’s Hospital of Los Angeles and Keck School of Medicine University of Southern California, Los Angeles, CA; Department of Radiology, Children’s Hospital of Los Angeles and Keck School of Medicine University of Southern California, Los Angeles, CA; Department of Critical Care Medicine, University of Pittsburgh, Pittsburgh, PA; Department of Radiology, Texas Children’s Hospital, Houston, TX; Department of Neurology, Children’s Hospital of Philadelphia, Philadelphia, PA; Department of Surgery, Children’s Hospital of Philadelphia, Philadelphia, PA; Department of Anesthesiology, Texas Children’s Hospital, Houston, TX

**Author notes:** Corresponding Author: Rafael Ceschin, Ph.D., Department of Radiology, Children’s Hospital of Pittsburgh of UPMC, 4401 Penn Ave., Pittsburgh, PA 15224. co-senior authorship (Ashok Panigrahy, Rafael Ceschin).

**Keywords:** Diffusion Tensor Imaging, Neonatal Imaging, Congenital Heart Disease

## Abstract

Advanced brain imaging of neonatal macrostructure and microstructure, which has prognosticating importance, is more frequently being incorporated into multi-center trials of neonatal neuroprotection. Multicenter neuroimaging studies, designed to overcome small sample sized clinical cohorts, are essential but lead to increased technical variability. Few harmonization techniques have been developed for neonatal brain microstructural (diffusion tensor) analysis. The work presented here aims to remedy two common problems that exist with the current state of the art approaches: 1) variance in scanner and protocol in data collection can limit the researcher’s ability to harmonize data acquired under different conditions or using different clinical populations. 2) The general lack of objective guidelines for dealing with anatomically abnormal anatomy and pathology. Often, subjects are excluded due to subjective criteria, or due to pathology that could be informative to the final analysis, leading to the loss of reproducibility and statistical power. This proves to be a barrier in the analysis of large multi-center studies and is a particularly salient problem given the relative scarcity of neonatal imaging data. We provide an objective, data-driven, and semi-automated neonatal processing pipeline designed to harmonize compartmentalized variant data acquired under different parameters. This is done by first implementing a search space reduction step of extracting the along-tract diffusivity values along each tract of interest, rather than performing whole-brain harmonization. This is followed by a data-driven outlier detection step, with the purpose of removing unwanted noise and outliers from the final harmonization. We then use an empirical Bayes harmonization algorithm performed at the along-tract level, with the output being a lower dimensional space but still spatially informative. After applying our pipeline to this large multi-site dataset of neonates and infants with congenital heart disease (n= 398 subjects recruited across 4 centers, with a total of n=763 MRI pre-operative/post-operative time points), we show that infants with single ventricle cardiac physiology demonstrate greater white matter microstructural alterations compared to infants with bi-ventricular heart disease, supporting what has previously been shown in literature. Our method is an open-source pipeline for delineating white matter tracts in subject space but provides the necessary modular components for performing atlas space analysis. As such, we validate and introduce Diffusion Imaging of Neonates by Group Organization (DINGO), a high-level, semi-automated framework that can facilitate harmonization of subject-space tractography generated from diffusion tensor imaging acquired across varying scanners, institutions, and clinical populations. Datasets acquired using varying protocols or cohorts are compartmentalized into subsets, where a cohort-specific template is generated, allowing for the propagation of the tractography mask set with higher spatial specificity. Taken together, this pipeline can reduce multi-scanner technical variability which can confound important biological variability in relation to neonatal brain microstructure.

## Introduction

Neonatal imaging studies are significantly more challenging than comparable adolescent and adult studies. Overall head size, rapidly developing structures in a short period of time, reduced control of motion in subjects, and higher water content in the brain tissue all contribute to the challenges in advancing neonatal imaging studies to equivalent adult standards.^1^ Imaging studies in disease populations are further hindered by low incidence rate, recruitment challenges, and data acquisition problems, which can all be exacerbated by abnormal anatomy. Thus, sufficiently large landmark neonatal studies more frequently necessitate the use of cross-scanner and cross-institutionally acquired imaging to be sufficiently powered to detect small effect sizes. This creates the additional problem of data harmonization. Latent vendor and scanner specific variances can introduce bias into cohort studies, decreasing statistical power and limiting the generalizability of the results.^2–4^

Here, we will delineate a diffusion imaging pipeline that is able to harmonize heterogeneously acquired neonatal imaging by hierarchically compartmentalizing the unwanted sources of variance, allowing for a final harmonization of multi-site, high anatomical variance data. The motivation behind this work is not to develop yet another standard for general neonatal diffusion imaging analysis, but rather to cover a need specific to multi-center studies using populations with pathological conditions. In these studies, state-of-the-art methods are insufficiently flexible when dealing with protocol heterogeneity, varying acquisition site and scanner, and abnormal anatomy. We believe this is of particular importance in identifying developing white matter neuroimaging biomarkers derived from multi-center neonatal neuroprotection trials. In this work we use as validation a large-scale dataset of neonates born with congenital heart disease (CHD) acquired at four different institutions. Congenital heart disease (CHD) is one of the most common birth defects, affecting nearly 1% of all births.^5,6^ Through advancements in surgical intervention, neonates born with CHD now have excellent survival rates. However, this population remains at a significant risk for more subtle cognitive-behavioral executive-functional deficits later in life, with a predilection for executive-functional disorders.^7–12^ Adolescents born with CHD are at risk for a constellation of behavioral and cognitive impairments, including ADHD, autism, and executive function disorders.^13,14^ The mechanism of these deficits requires further elucidation, and prognostic biomarkers are needed to identify these deficits at the earliest possible time in development.

Diffusion tensor imaging (DTI) is an advanced MR technique that models microstructural white matter connections throughout the brain. DTI has been used extensively to map white matter connections in normal populations and a plethora of disease subtypes, including autism, dementia, and trauma.^15–18^ Moreover, DTI measures provide an effective estimation of myelination and overall white matter development in neonatal imaging. DTI has increasingly been incorporated, both in clinical trials and routine diagnostic workflows, as a complement to surgical planning and injury characterization, but primarily in adult populations. While analysis and clinical methods in adult imaging have matured into robust, reproducible standards, comparable methods for neonatal imaging are still lagging.

Existing DTI analysis techniques in neonates require a trade-off between labor intensive, but precise individual-level methods and more limited, but faster automated group-level approaches.^19–27^ Manually delineated tracts in native subject space are considered the gold standard of DTI tractography. This approach requires the user to manually draw regions of interest (ROI) and regions of avoidance (ROA) in each subject’s diffusion image, for each tract of interest. Manual delineation provides the most accurate tract outputs, as each region is specifically placed with consideration to each subject’s variable anatomical features.^28^ However, this method is prohibitively time consuming when analyzing large datasets. Manual tractography can be prone to user bias in both mask generation and often-used “tract pruning” steps, resulting in diminished reproducibility. Moreover, native space tractography lacks inter-subject correspondence between tracts, as each subject tract has its own unique curvature and volume. Relative position can be modeled across subjects, but with lower reproducibility and anatomical accuracy. As such, purely native space analyses are limited to global tract metrics such as average fractional anisotropy or other derived diffusivity metrics, resulting in relatively low biomarker granularity, despite the required manual labor. ^28,29^

The work presented here aims to remedy two common problems that exist with the methods mentioned so far: 1) The general lack of objective guidelines for dealing with anatomically abnormal anatomy and pathology. Often, subjects are excluded due to subjective criteria, or due to pathology that could be informative to the final analysis. This leads to the loss of reproducibility and statistical power. 2) Variance in scanner and protocol in data collection can limit the researcher’s ability to harmonize data acquired under different conditions or using different clinical populations. This proves to be a barrier in the analysis of large multi-centered studies and is a particularly salient problem given the relative scarcity of neonatal imaging data. We provide an objective and data-driven semi-automated neonatal processing pipeline designed to harmonize compartmentalized variance data acquired under different parameters. This is done by first implementing a search space reduction step of extracting the along-tract diffusivity values along each tract of interest, rather than performing whole-brain harmonization. This is followed by a data-driven outlier detection step, with the purpose of removing unwanted noise and outliers from the final harmonization. We then use an empirical Bayes harmonization algorithm performed at the along-tract level, with the output being a still spatially informative, yet lower-dimensional space. We test the null hypothesis that after harmonization. There should be no observed differences across similarly stratified subjects, while retaining the across-group variances. After applying our pipeline to this large multi-site dataset of neonates and infants with CHD, we show that infants with single ventricle cardiac physiology demonstrate greater differences in developing white matter microstructural alterations compared to infants with bi-ventricular heart disease, which has been shown by previous literature.^30,31^ Our method is an open-source pipeline for delineating white matter tracts in subject space, but provides the necessary modular components for performing atlas space analysis. We minimize the dependency of a standard atlas by only using it to propagate robust masks, retaining subject space fidelity.

## Methods

### Subjects

A total of 652 near term born neonates (> 34 weeks GA) with congenital heart disease (with both single ventricle and bi-ventricular cardiac anatomy)were prospectively and retrospectively recruited at UPMC Children’s Hospital of Pittsburgh (CHP), Children’s Hospital of Philadelphia (ChoP), Children’s Hospital Los Angeles (CHLA), and Texas Children’s Hospital (TCH). Institutional Review Board approval was granted at each scanning site, and all imaging was acquired with written parental consent. Inclusion criteria for the study across all four sites was the presence of CHD (single or bi-ventricular physiology) and a neuroimaging study. Of note, these patients were recruited for different types of research studies with the purpose of correlating neuroimaging biomarkers with fetal imaging time points, concurrent post-natal neuro-diagnostic studies (NIRS/optical imaging/EEG) and correlated with neurodevelopmental outcomes. Multiple prior site-specific publications document the specific inclusion criteria.^32–35^ 221 infants were recruited from CHP (101 included in analysis), 225 from ChoP (129 included), 107 from CHLA (77 included), and 99 from TCH (89 included). Figure 1 shows a summary of the demographic distribution of subjects acquired from each site.

**Figure 1.**
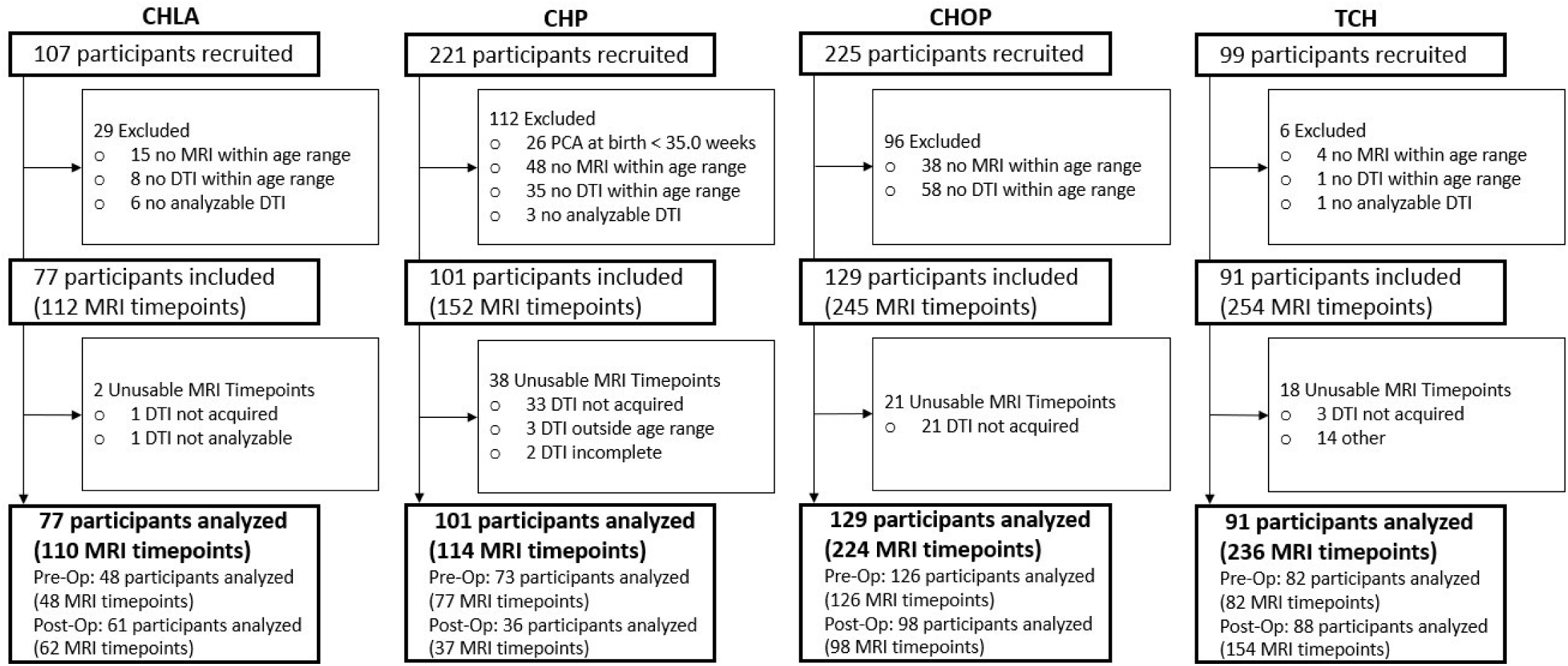
Summary of population for each cohort.

### MRI Acquisition

Infants were scanned at close to term equivalent age, or when deemed clinically stable. Infants recruited at CHP were scanned on a 3T Siemens Skyra (32-channel head coil, FOV=256mm, voxel dimensions=2×2×2mm, TE/TR=83/7800ms with 42 directions at B=1000s/mm^2^) or GE Signa HDxt (32-channel head coil with FOV=160-260mm, voxel dimensions 2×2×2.2-6mm with 30 directions at B=700-1000s/mm^2^). Infants imaged at TCH were scanned on a 1.5T Philips Achieva (8 channel head coil, FOV=200mm, voxel dimensions=2×2×2.7mm, TE/TR=90/6065ms, 15 directions at B=860s/mm^2^). Infants scanned at CHoP were scanned on a 1.5T Siemens Avanto (8 channel head coil, FOV=220mm, voxel dimensions=1.5×1.5×5.2mm, TE/TR=93/5600ms, 80 directions [20 unique] at B=1000s/mm^2^). Infants imaged at CHLA were scanned on a 3T Philips Achieva (8-channel head coil, FOV=190mm, voxel dimensions=2×2×2mm, TE/TR=74/8000 with 32 directions at B=700s/mm^2^). Not enough data was collected for four scans (of three infants) and two moved too much to allow reconstruction. There was a persistent spike artifact for one infant. For six infants there was data corruption before analysis could be conducted.

### Diffusion Imaging of Neonates by Group Organization (DINGO)

The overarching goal of this pipeline is to provide a semi-automated framework for the harmonization of subject-space tractography generated from diffusion tensor imaging acquired across varying scanners, institutions, and populations. **Figure 2** shows a high-level overview of the cohort-specific harmonization framework. Datasets acquired using varying protocols are compartmentalized into subsets, where a cohort-specific template is generated, allowing for the propagation of the tractography mask set with higher spatial specificity. To reduce variance of label propagation and minimize manual effort, only one set of labels are generated for the entire analysis, derived from a “master template” created from combining all dataset specific templates. The master mask set is propagated through each cohort specific template, resulting in group-specific sets of labels.

**Figure 2.**
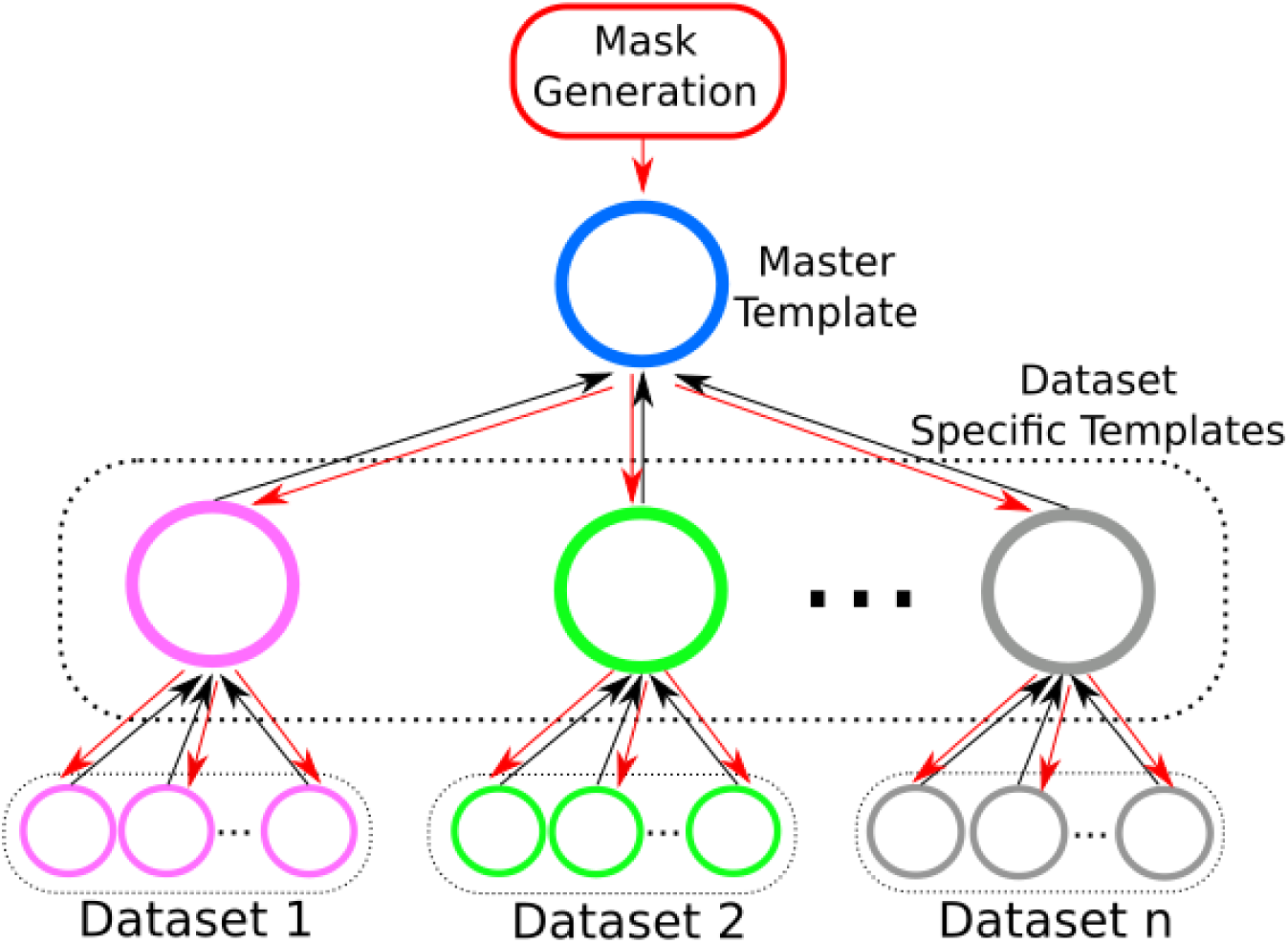
Cohort compartmentalization framework. Every participant in a cohort is registered to its most representative subject to generate a cohort specific mean FA map. Regions of interest/avoidance are created in this mean FA space and transformed back into individual space for fiber tracking. Red arrows show the propagation of the masks from template space into subject space, black arrows show subject images propagated into template space.

**Figure 3** shows a detailed view of the group-level workflow of the proposed pipeline, named Diffusion Imaging of Neonates by Group Organization (DINGO). Detailed descriptions of each step will be provided below. DINGO first generates a cohort-specific template by iteratively co-registering all subjects in the cohort, generating a group-wise average atlas. This atlas is used for the generation of ROI and ROA for the desired tracts. Here, we will refer to the set of ROIs and ROAs for each tract as the Mask Set. The mask sets are propagated onto each subject’s native DTI space for fiber tracking. At each iteration of the previous steps, quality control is performed by carefully chosen criteria: spatial maps, along tract metrics, and group diffusion metrics. If necessary, mask refinement steps are performed to improve these metrics until no significant improvement is observed. This is described in detail below. The final step is to analytically find outliers, and if deemed appropriate, to remove them from the group, followed by a final pass through the workflow. The identified outliers are then grouped into their own sub-cohort, where a new cohort-specific template is generated and processed through DINGO.

**Figure 3.**
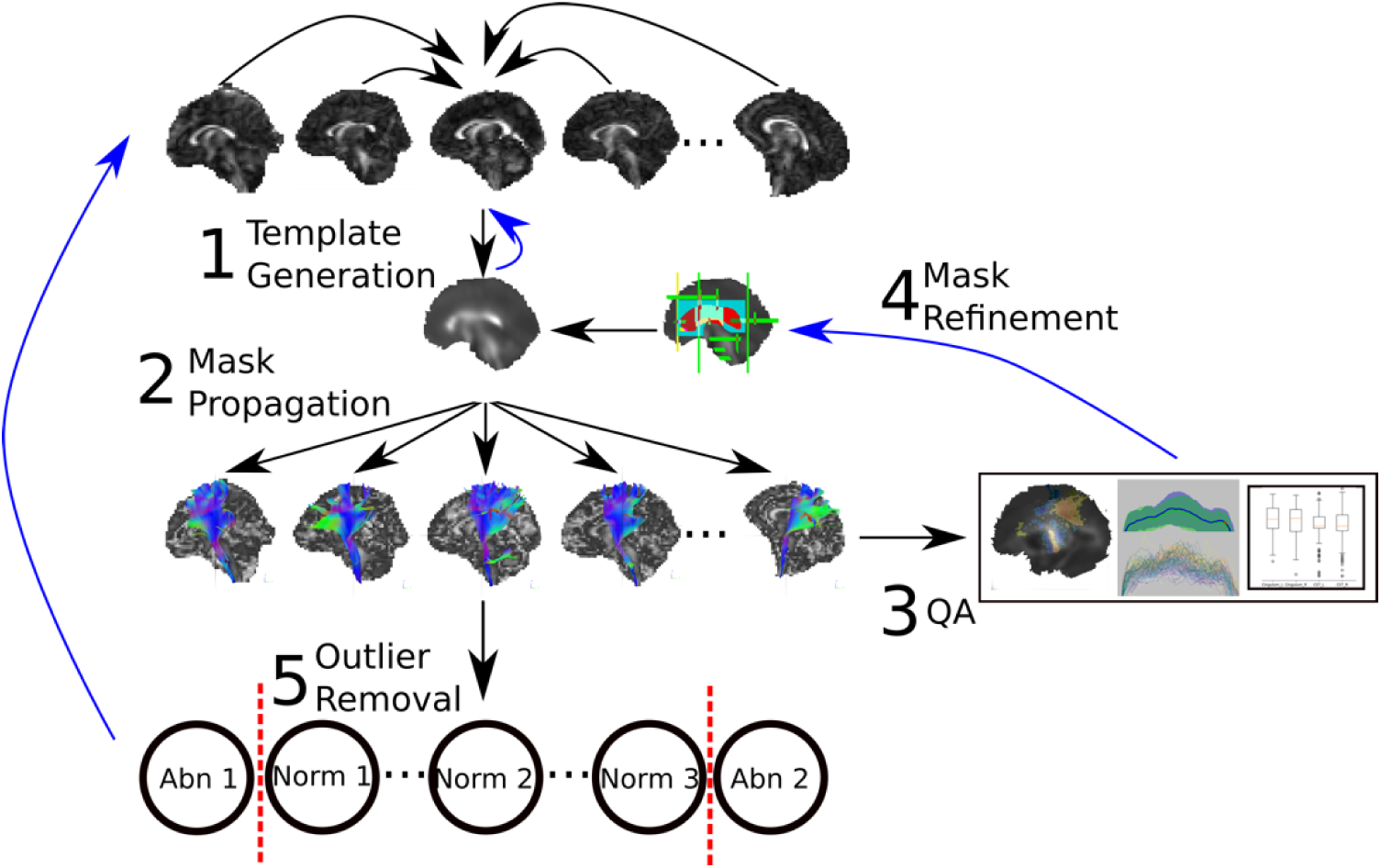
DTI Processing Workflow. ROI/A masks created in template space are warped to individual space for tractography with DSI Studio. Generated tracts are evaluated against a subset of manually delineated subject-space masks on the basis of DICE coefficients (spatial overlap), tract averaged statistics, and along tract measures to create satisfactory group masks. Outliers are identified via isolation forest and either removed from analysis or grouped into a new sub-cohort if sufficient outliers with similar spatial similarity are identified.

The core framework of the DINGO pipeline is built using the Nipype library.^36^ Nipype is an open source pipeline development library developed to promote open and reproducible neuroimaging workflows, providing direct interfaces to standard neuroimaging tools, including FSL^37^ and DSI Studio. ^38^The full DINGO source code can be found at https://github.com/PIRCImagingTools/DINGO.git.

### Pre-Processing

First, each subject’s diffusion tensor images are inspected for motion artifacts, and gradient volumes with strong artifacts are removed. Images have the brain extracted using FSL Brain Extraction Tool (BET),^39^ then are eddy current and motion corrected using the standard DSI studio workflow. The estimated motion parameters are applied to the diffusion vectors prior to tensor reconstruction. Fractional anisotropy (FA), radial diffusivity (RD), axial diffusivity (AD), and mean diffusivity (MD) maps are generated for each subject in native space and the FA serve as the input for the template generation step.

### Template Generation

The template generation step of DINGO leverages the existing cohort-specific template creation workflow developed by the existing Tract-Based Spatial Statistics (TBSS) method.^40,41^ Each subject within the cohort is registered to every other subject, and the subject with the smallest average deformation to every other subject is chosen as the “representative subject” of the cohort. This becomes the template space for group registration. Each subject is then non-linearly transformed into this space, and an average FA map is generated for the cohort. The non-linear transformation is performed using the Advanced Normalization Tools (ANTS).^42^ This FA map then becomes the new target of registration, and all subjects are again transformed into this space, generating a new average FA map.

### Iterative Mask Set Refinement

Barring post-mortem dissection, there is no objective anatomical gold-standard for imaging based tractography. Manual approaches use anatomy and segmentation experts to evaluate the accuracy of the tracts, including prior knowledge-guided “pruning” of spurious connections. Therefore, to measure the accuracy of our iteratively developed semi-automated method, we first needed to generate a “gold-standard” set by manually delineating the mask sets for the desired tracts. The final evaluation of the accuracy of the automated method uses a combination of quantitative comparisons to this “gold-standard” as well as expert-guided interpretation. We delineated the following tracts: Genu, Body, and Splenium of the Corpus Callosum; anterior and posterior segments of the superior longitudinal fasciculus (SLFA and SLFP, respectively); Inferior Longitudinal Fasciculus (ILF), Fronto-Occipital Fasciculus (FOF), Cingulum, and Cortical Spinal Tract (CST). Manual mask set delineation was performed following the guidelines published by Fernandez-Miranda et al.^43^ **Table 1** shows the ROIs and ROAs comprising each mask set, and visualization of each manual mask set is provided as supplemental material. All subjects from the CHP and CHLA datasets were manually delineated.

**Table 1.** Patient Demographics

| Demographic Variables | ALL | CHLA | CHP | CHOP | TCH |
| --- | --- | --- | --- | --- | --- |
|  | % | % | % | % | % |
| Male | 59.19 | 61.84 | 71.29 | 50.39 | 56.04 |
| Late Preterm* | 2.76 | 3.90 | 0.99 | 0.78 | 6.59 |
| Single Ventricle | 45.59 | 61.84 | 36.63 | 41.09 | 48.35 |
| Age at MRI | M (SD) | M (SD) | M (SD) | M (SD) | M (SD) |
| Mean PNA at Pre-Op MRI - weeks | 1.5 (7.81) | 0.5 (1.04) | 4.1 (15.98) | 0.5 (0.36) | 1.2 (0.72) |
| Mean PCA at Pre-Op MRI - weeks | 40.3 (7.95) | 39.0 (1.95) | 43.1 (16.06) | 39.4 (0.99) | 39.8 (1.64) |
| Mean PNA at Post-Op MRI - weeks | 8.2 (11.84) | 6.0 (3.77) | 13.2 (23.34) | 1.6 (0.57) | 12.2 (11.64) |
| Mean PCA at Post-Op MRI - weeks | 46.9 (11.84) | 44.8 (3.71) | 52.1 (23.56) | 40.5 (1.04) | 50.6 (11.64) |

\*Notes: Late Preterm = born 35.0 – 36.9 weeks PCA

The first iteration of the pipeline was performed by duplicating the above mask sets, following the identical anatomical guidelines, on the generated cohort template. The masks were then propagated into each subject’s native diffusion space using the previously calculated non-linear transforms. Each tract was delineated in DSI studio using a deterministic tracking algorithm an FA threshold of 0.1 and angular threshold of 45 degrees with no manual pruning. We used four increasingly granular metrics to measure the accuracy of the semi-automated approach. At each successive mask-refinement iteration, more emphasis is placed on the more granular measure. First, as a qualitative measure of cohort-level accuracy, we projected both the manually delineated tracts and automated tracts onto the cohort-specific atlas, displaying the spatial distribution of each tract and level of agreement. This allows for the detection of obvious points of failure in the pipeline, such as consistently present spurious fibers or anatomically incongruent delineations, as well as a general overview of the variance in anatomical tract location.

We then quantitatively measured the degree of spatial overlap between each manually delineated tract and gold standard tract using the Dice Similarity Coefficient (DSC).^44^ Each tract’s mask set is then independently refined at the master template to improve this measure prior to the next iteration. The directive of mask refinement is set to alter the masks to make them more robust to mis-registration errors, while controlling for the appearance of spurious tracts. The third measure used was the distribution of the mean fractional anisotropy of each tract. This provides a quantitative measure of the whole tract and can indicate slight global mis-registration or dilation of the tract body that may lead to decreased anisotropy values.

Finally, the final metric used to quantitate the accuracy of the automated method was using along-tract analysis. Along-tract analysis measures the mean anisotropy of each tract at each point in its principal orientation. This approach measures variations at local points in each tract and can be indicative of regional failures in the mask set delineation. **Figure 4** shows the along tract FA measure comparing the manually delineated and the automated generation of each tract. We see no statistically significant differences in tract FA, however, we do observe non-significant, higher variance in regions of low FA and at tract extremes. This method is informative of regions of low reliability in tract delineation and is an objective measure of regions that should be excluded from comparative analyses between groups.

**Figure 4.**
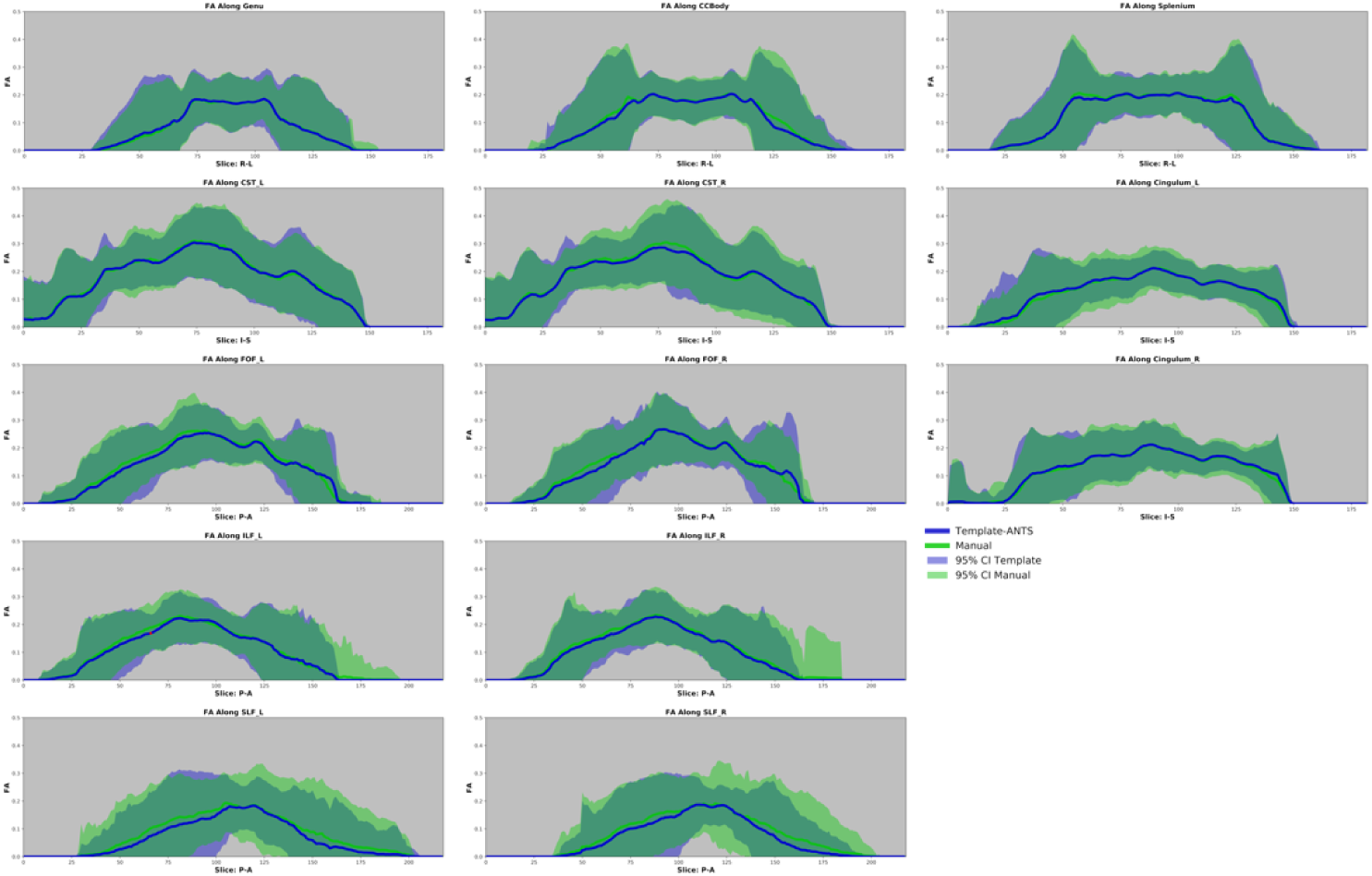
Spatial Distribution of Manual vs. Automated Along-Tract FA. Comparison of template generated tracts and manually delineated (in subject space) tracts. FA is measured along a tract’s primary direction, and statistical comparison is done at each voxel using permutation. No statistical significance was observed between manually delineated tracts and templated generated tracts after three iterations of mask refinement.

### Outlier Detection

Prior to the final harmonization step, we run an outlier detection algorithm to identify significantly abnormal subjects within each group. This acts as an automated data check to alert users of potential technical problems, as well as significantly abnormal subjects, that may have cleared preliminary QC. We place this processing step prior to harmonization, as harmonization methods can be sensitive to outliers, which could lead to biased correction factors.^2,3,45^ Several approaches for outlier detection were explored, including regression, principal component analysis (PCA), and PCA combined with support vector machines (SVM). We chose to implement an Isolation Forest (IF) algorithm, as it is a computationally cheap, data agnostic approach to outlier selection, and less prone to overfitting compared to SVMs.^46^ Isolation Forests, as its name implies, takes advantage of the binary tree structure inherent to Random Forest classification algorithm. IF trains a random forest algorithm on the entire dataset (all concatenated along-tract FA values) and uses the average path length of each subject’s set of decision trees to determine how likely it is to be an outlier. The general principle is that a subject with a consistently short path length across all trees is more likely to be an outlier, as it is readily separated from the remainder of its cohort. This method is intended to identify subtle technical or biological outliers that may have cleared preliminary quality control, as it is applied to the already delineated along-tract measures. We used the ScikitLearn^47^ implementation using the recommended default hyperparameters and 300 estimators.

### Data Harmonization

In the setting of retrospectively acquired data, we are limited in the techniques we can apply to harmonize site parameters – retroactively scanning phantoms is generally not feasible and may not be representative of the scanner profile at the time of study. Bayesian approaches to data harmonization provide a flexible set of tools that incorporate the inherent uncertainty of the estimated parameters into the analysis. Empirical Bayes methods, including the recently published ComBat approach,^48^ estimate from the input data a set of scanner-specific correction factors that are dependent on the covariates of interest. This approach tries to estimate the distribution that contributes most to the covariates being analyzed, while shrinking away the variance contributed by external factors (including inter-scanner variability). DINGO performs an empirical Bayes correction at the along-tract level using the publicly available NeuroCombat package^45^. This generates a new harmonized diffusion metric (FA, MD, RD, or AD) at each point along the tract of interest.

### Statistical Analysis and Evaluation

We evaluated the performance of DINGO on four independent datasets of neonates with CHD acquired at different institutions. As each cohort comes from populations of matched cardiac pathologies, we expect to observe similar diffusion metric distribution between each group after harmonization using ComBat. We tested the hypothesis that neonates born with a single ventricle heart defect (SV) would show reduced anisotropy (FA) and increased diffusivity (mean, radial and axial diffusivity) compared to bi-ventricular heart defects in developmentally vulnerable white matter tracts. Along-tract statistical significance was assessed on post-menstrual age-adjusted diffusivity metrics (FA,RD,MD,AD) at pre- and post-surgical intervention time points. P-values were calculated using re-randomization tests (1000 permutations per tract slice), for each tract, using Welch’s T-statistic for unequal variance. Multiple comparison correction was done using the stepdown max-T algorithm^49^.

## Results

### Subjects

Of the 592 infants recruited from CHP, 101 were included in the final analysis. 70 imaging time points at CHP were acquired prior to first surgical intervention, with 36 scanned post-surgery. Of the 107 infants recruited from CHLA, 77 were included. 49 time points were acquired pre-surgical intervention, and 61 imaging time points were post-surgery. Of the 225 infants recruited from CHoP, 129 were included in the analysis. 124 time points scanned pre-intervention, and 98 scanned post-intervention were included. Of the 99 infants recruited from TCH, 91 were included in the final analysis. 82 time points scanned pre-surgical intervention were included, with 83 scanned post-intervention. For final comparison between single vs. biventricle outcomes, subjects with GA below 34 were excluded. Pre-op time points were restricted to < 6 days post-natal age, and post-op time points were restricted to < 25 days.

### Template Generation

The chosen neonate to initialize the template for the CHP cohort was born at 39.1 weeks gestation, scanned at 0.4 weeks post-natal, pre-/post-surgical [diagnosis: double outlet right ventricle with hypoplastic left ventricle and heterotaxy]. The chosen infant for the TCH cohort was born at 38.1 weeks gestation, scanned at 0.9, 1.9 weeks post-natal, pre-,post-surgical [HLHS]. The representative infant for the CHoP cohort was born at 39.0 weeks gestation, scanned at 0.3, 9.0 weeks post-natal, pre-,post-surgical intervention [d-TGA]. The chosen infant for the CHLA cohort was born at 39.0 weeks gestation, scanned at 0, 9 weeks post-natal. Supplemental Figure 1 shows each site’s selected representative subject, and the final generated atlas.

### Iterative Mask Set Refinement

**Figure 5** shows the DICE coefficients between the gold-standard manually delineated tracts and the automatically generated tracts after final iteration. Tracts with higher average FA (CST and CC fibers) showed the highest DICE coefficients compared to manual delineated fibers, while peripheral cortico-association fibers (SLF) showed the lowest DICE coefficients. Supplementary figure 2 shows a comparison between mask sets for each tract pre- and post-refinement. In total, 4 iterations were required before reaching significantly diminishing returns in DICE coefficients for all tracts. Masks were altered to be more robust to misregistration errors, generally requiring larger coverage for each mask, and additional exclusion masks to delineate the search space for fiber tracking.

**Figure 5.**
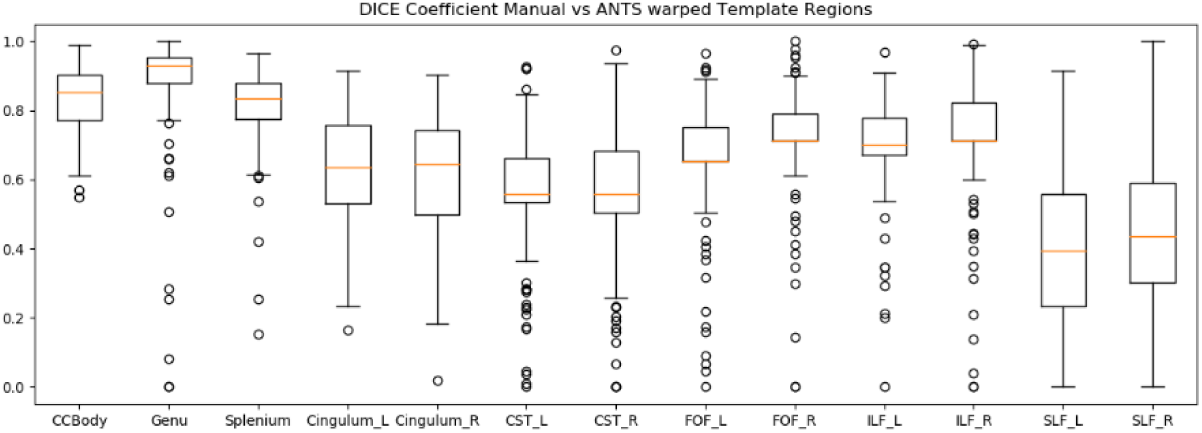
DICE coefficient by tract comparing those generated with warped cohort masks and individual masks. Tracts with higher average FA (CST and CC fibers) showed the highest DICE coefficients compared to manual delineated fibers, while peripheral cortico-association fibers (SLF) showed the lowest DICE coefficients

### Outlier Detection

A total of 8 (3 pre-op, 5 post-op) subjects were identified by the Isolation Forest algorithm as outliers. Figure 6 shows an example of inliers compared to outliers detected by IF. As the IF algorithm uses all concatenated tracts to detect outliers, we do not attribute the selection criteria to individual tracts. However, it is useful to visualize each individual tract as a diagnostic step. Qualitative interpretation of outlier selection shows that outliers were selected for two primary reasons: 1) low signal resulting in fewer fiber tracts per tract of interest, and 2) spurious tracts causing missing or excessive bundles outside the expected anatomical location of the desired tract. Supplemental figure 3 shows the effect of including the selected outliers in the final harmonization step, in the left cortico-spinal tract at post-operative scan. Despite only including an additional 5 subjects, a noticeable increase in cross-site variance can be seen, particularly in areas of lower signal such as crossing fibers.

**Figure 6.**
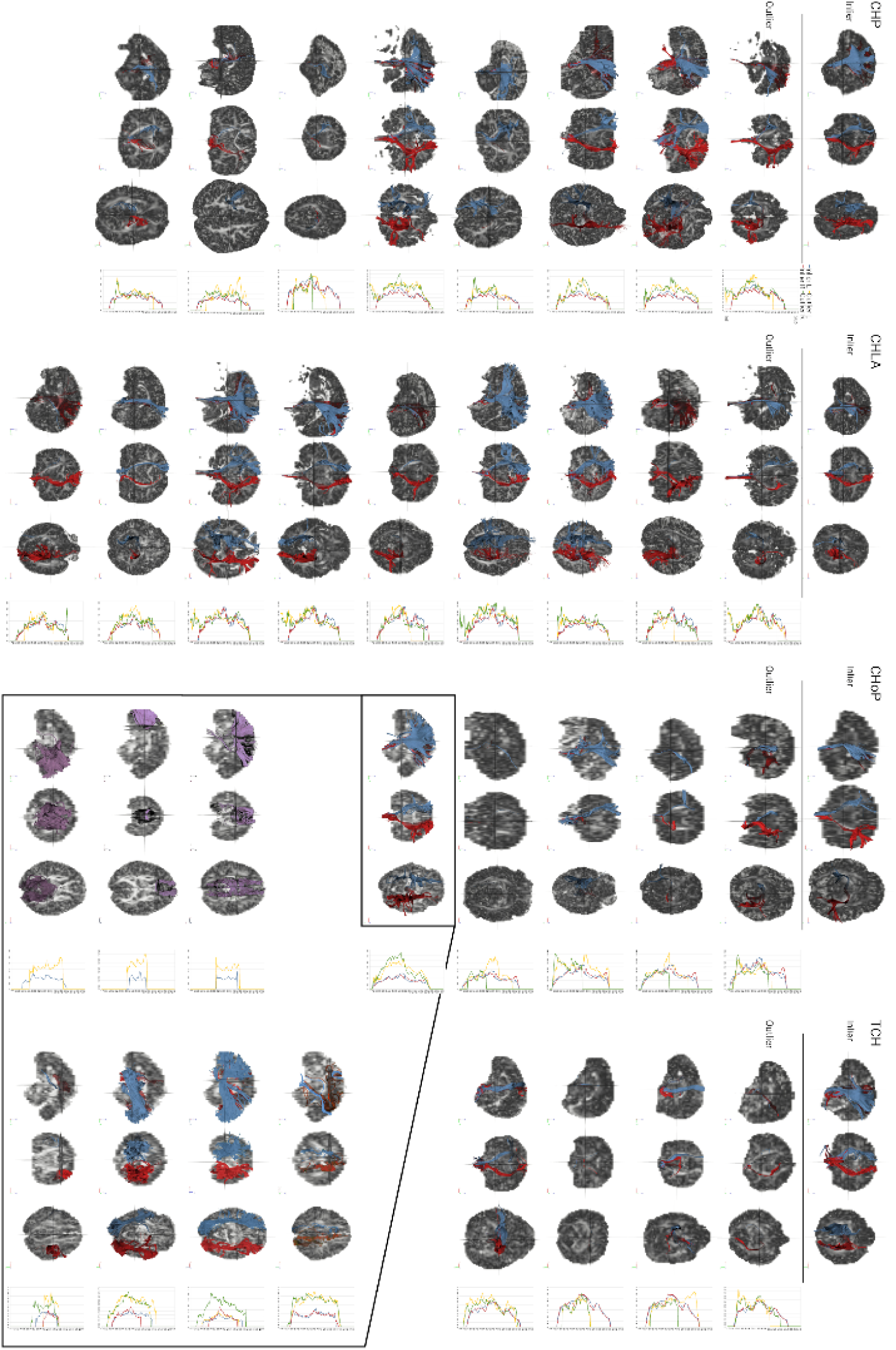
Comparison of outliers identified by along tract isolation forest and most representative individual per cohort for CST, both 3D and along tract. Isolation Forest aggregates all tract values as its input, and selects outliers based on overall deviance from the population across all tracts. Breakout box shows all tracts for a single subject acquired at CHoP. While the CST appears visually normal, the subject was selected as an outlier as a result of asymmetrical delineation of peripheral tracts, notedly right SLF and bilateral ILF.

### Cross-Site Harmonization

We applied ComBat independently to the pre-op timepoint scans and post-op, harmonizing the cross-site variance across scans. Covariates included in the harmonization were each infant’s post-natal age at time of scan, gestational age, and single-/bi-ventricle surgical repair. **Figure 7** shows the observed effect size for the left cortico-spinal tract pre- and post-harmonization at post-surgical intervention. Note the cross-site variance prior to harmonization is larger in regions of lower signal (lower FA).

**Figure 7.**
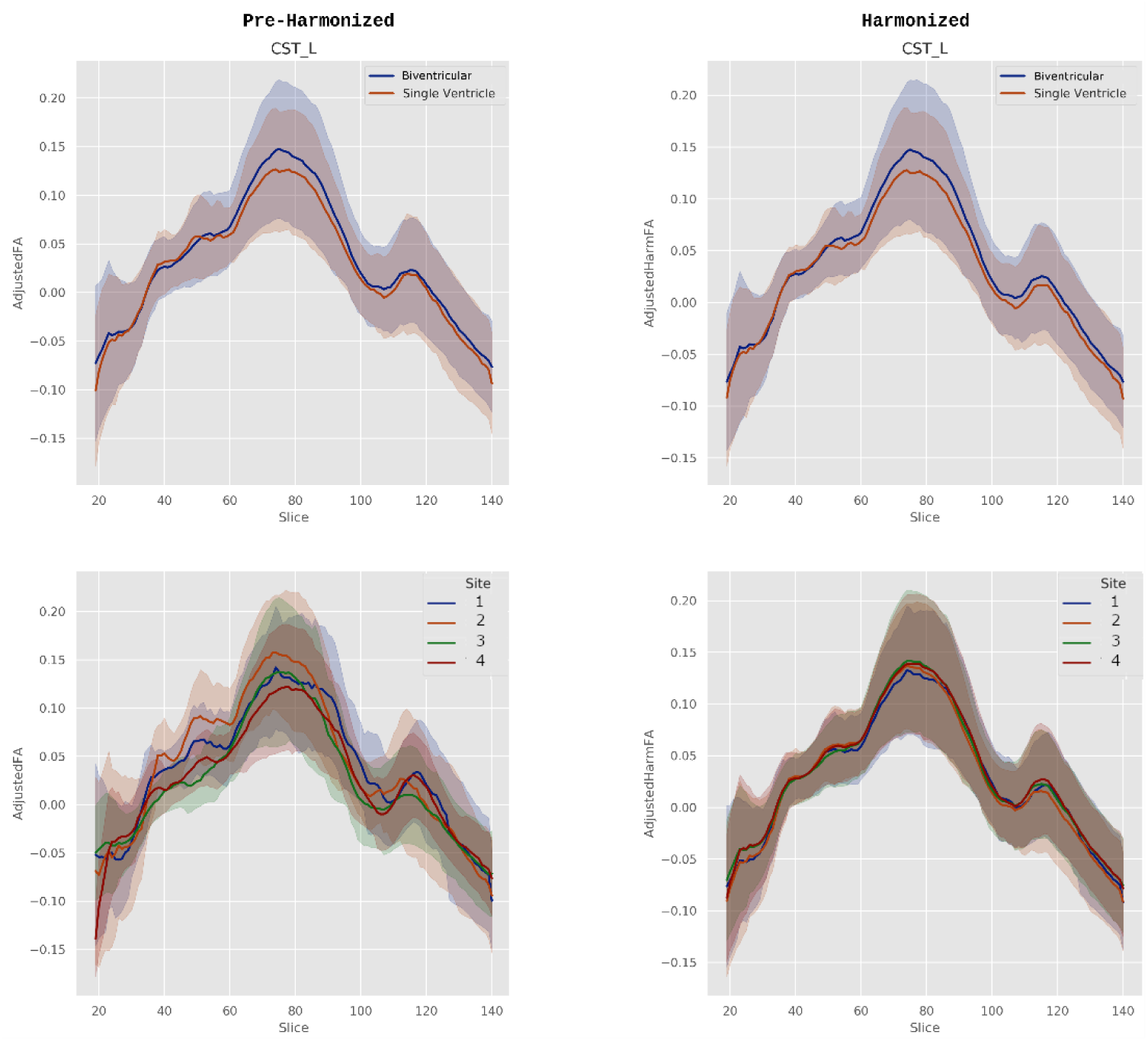
Raw and ComBat harmonized left corticospinal tract age-adjusted FA in neonates with single or biventricular repair acquired across four institutions. Top panels compare patients with single-vs. Biventricular reconstructions. Bottom shows the same subjects grouped by site. Left shows pre-harmonized FA values, right panels are post-harmonization FA. Note that the cross-site variance is larger than the biological variance prior to harmonization.

### Single vs. Bi-Ventricle Differences at Pre- and Post-Op

There was a statistically significant difference in FA (bi-ventricle reconstruction > single ventricle reconstruction) along the left FOF (**Figure 8**) and bilateral CSTs (**Figure 9**) in the pre-op time points. The remaining tracts analyzed did not show significant differences in FA at pre-op scan. In the post-op scan, we note statistically significant differences in FA (bi-ventricle > single ventricle) along bilateral CST (**Figure 10**), bilateral cingulum (**Figure 11**), bilateral FOF (**Figure 12**), and corpus callosum genu, splenium, and body (**Figure 13**). We did not see statistically significant differences in SLF at the post-operative timepoint.

**Figure 8.**
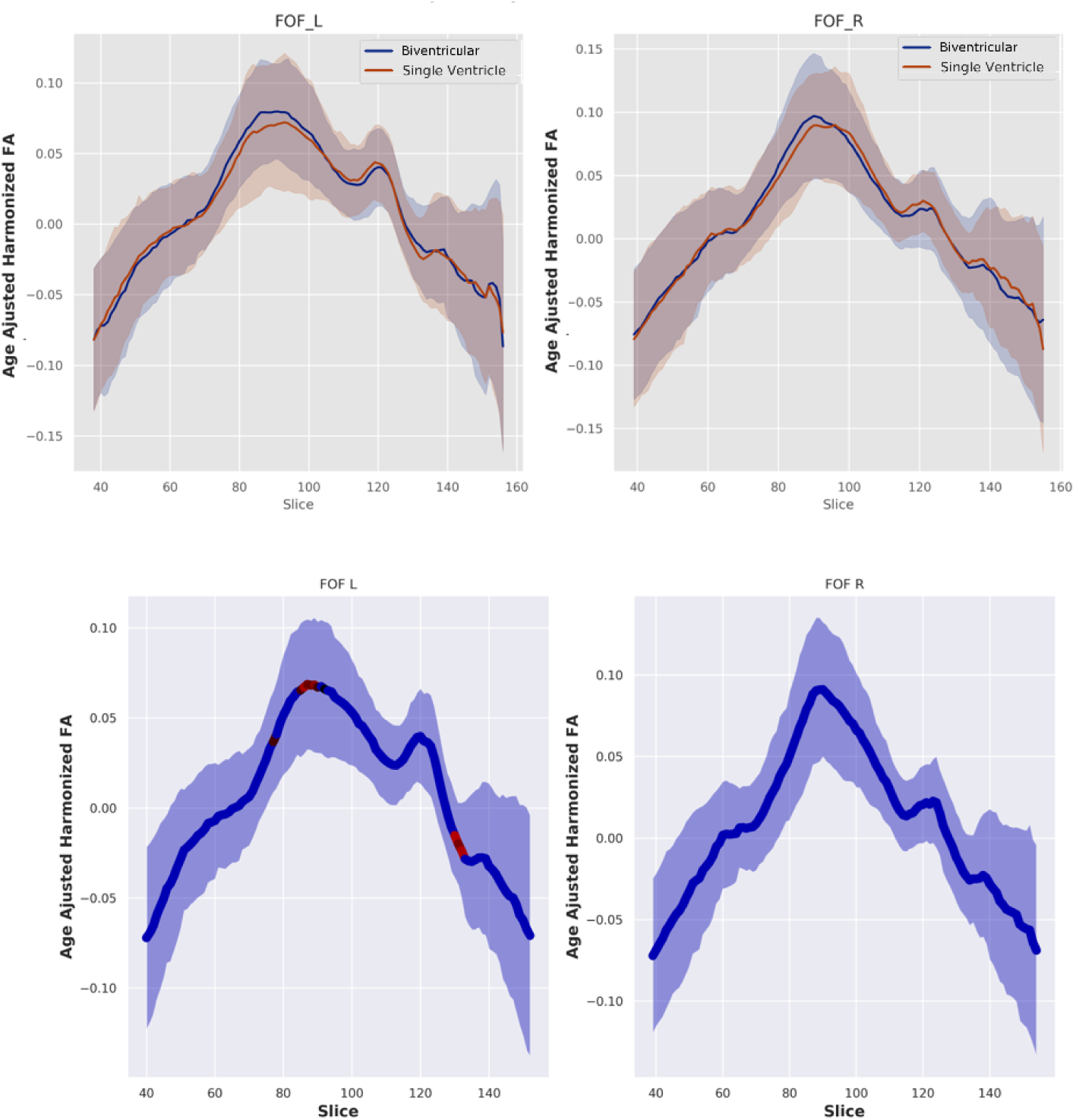
Pre-operative along tract age-adjusted FA that was harmonized with ComBat in Fronto-occipital Fasciculus (FOF). A) mean distribution between single vs. bi-ventricular repair groups. B) Statistically significant differences (p < 0.05) between single and biventricular repair projected onto mean along-tract FA in red.

**Figure 9.**
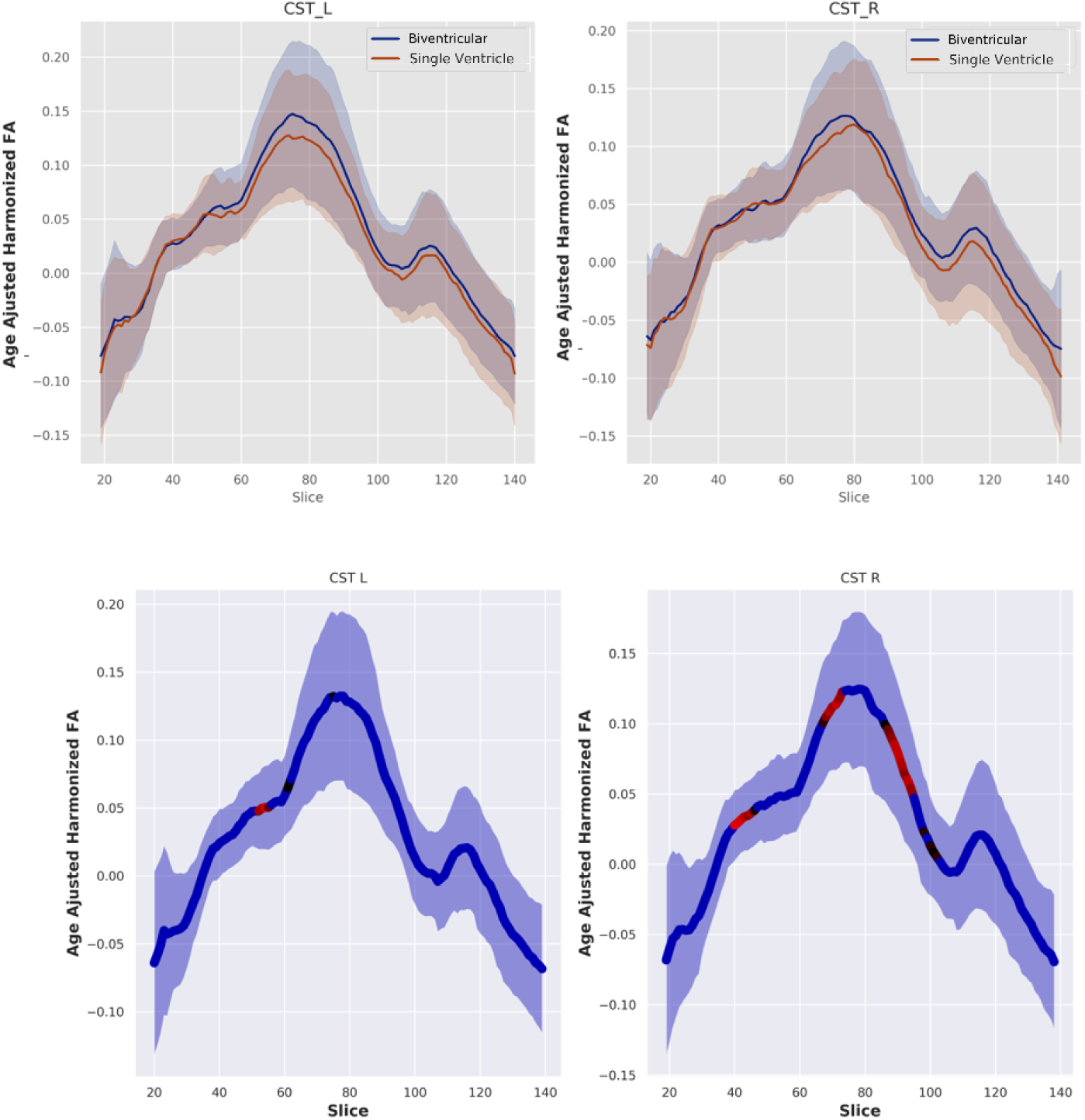
Pre-operative along tract age-adjusted FA that was harmonized with ComBat in Corticospinal tract (CST). A) mean distribution between single vs. bi-ventricular repair groups. B) Statistically significant differences (p < 0.05) between single and biventricular repair projected onto mean along-tract FA in red.

**Figure 10.**
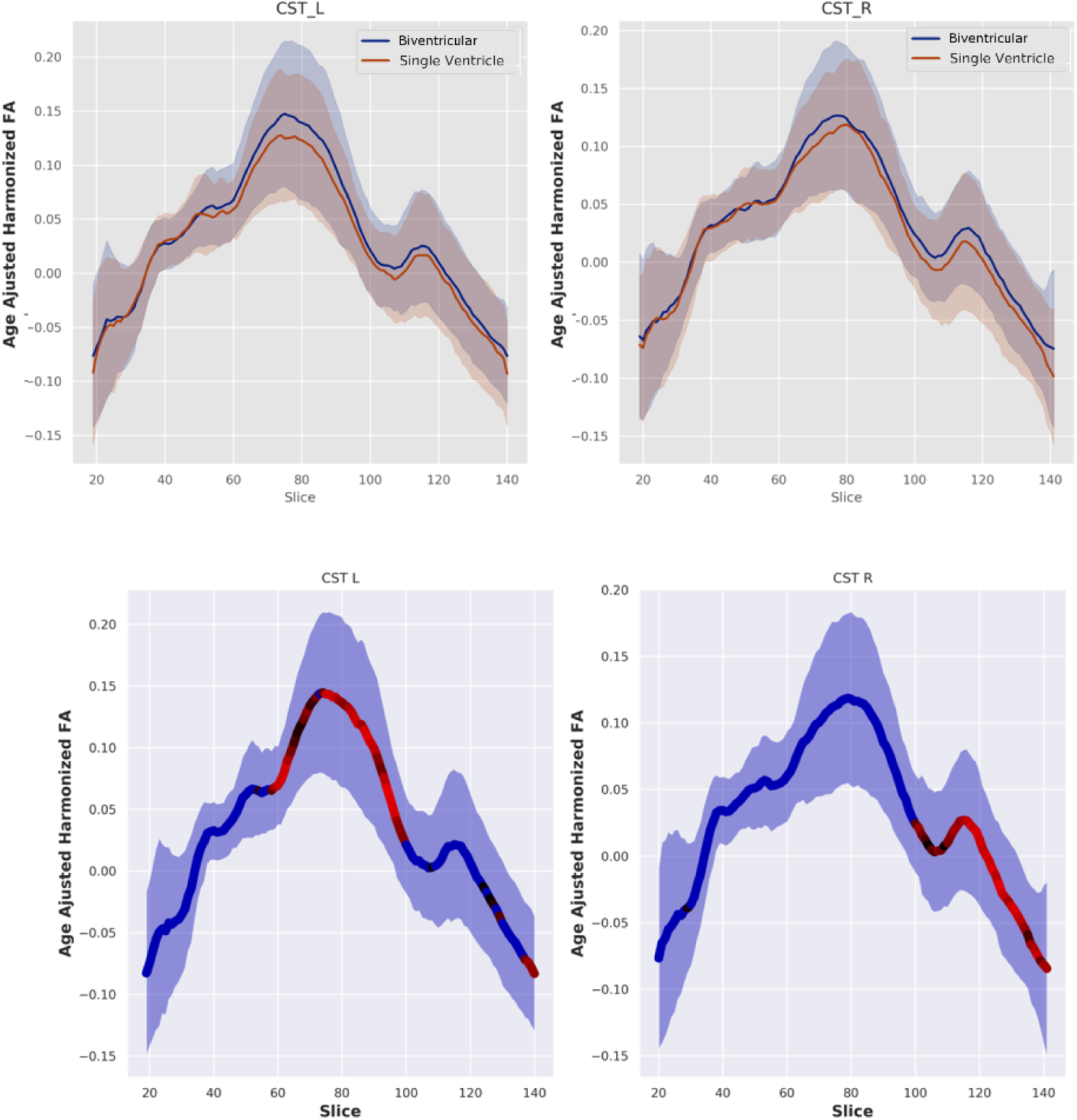
Post-operative along tract age-adjusted FA that was harmonized with ComBat in Corticospinal tract (CST). A) mean distribution between single vs. bi-ventricular repair groups. B) Statistically significant differences (p < 0.05) between single and biventricular repair projected onto mean along-tract FA in red.

**Figure 11.**
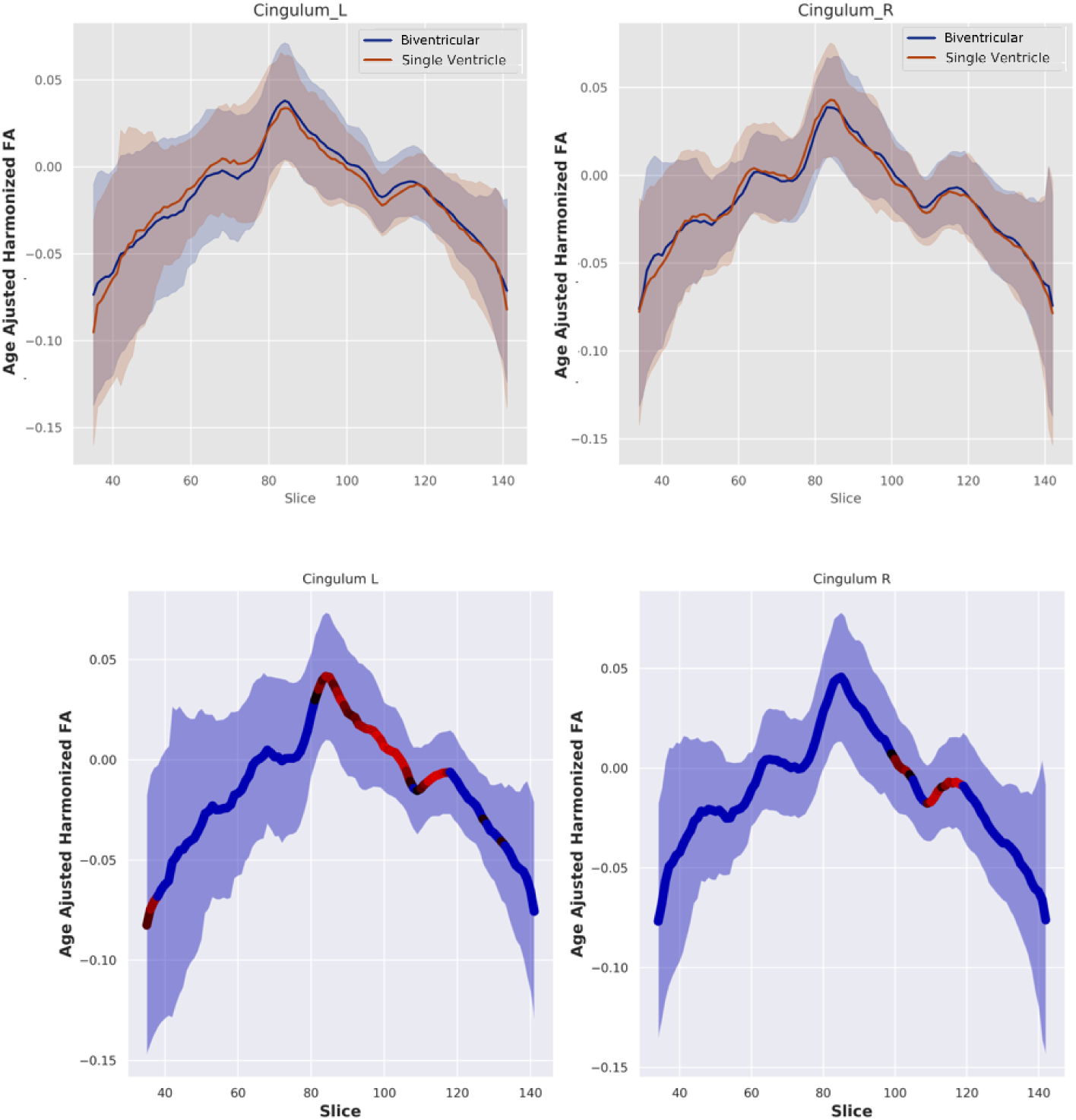
Post-operative along tract age-adjusted FA that was harmonized with ComBat in Cingulum.(A) mean distribution between single vs. bi-ventricular repair groups. B) Statistically significant differences (p < 0.05) between single and biventricular repair projected onto mean along-tract FA in red.

**Figure 12.**
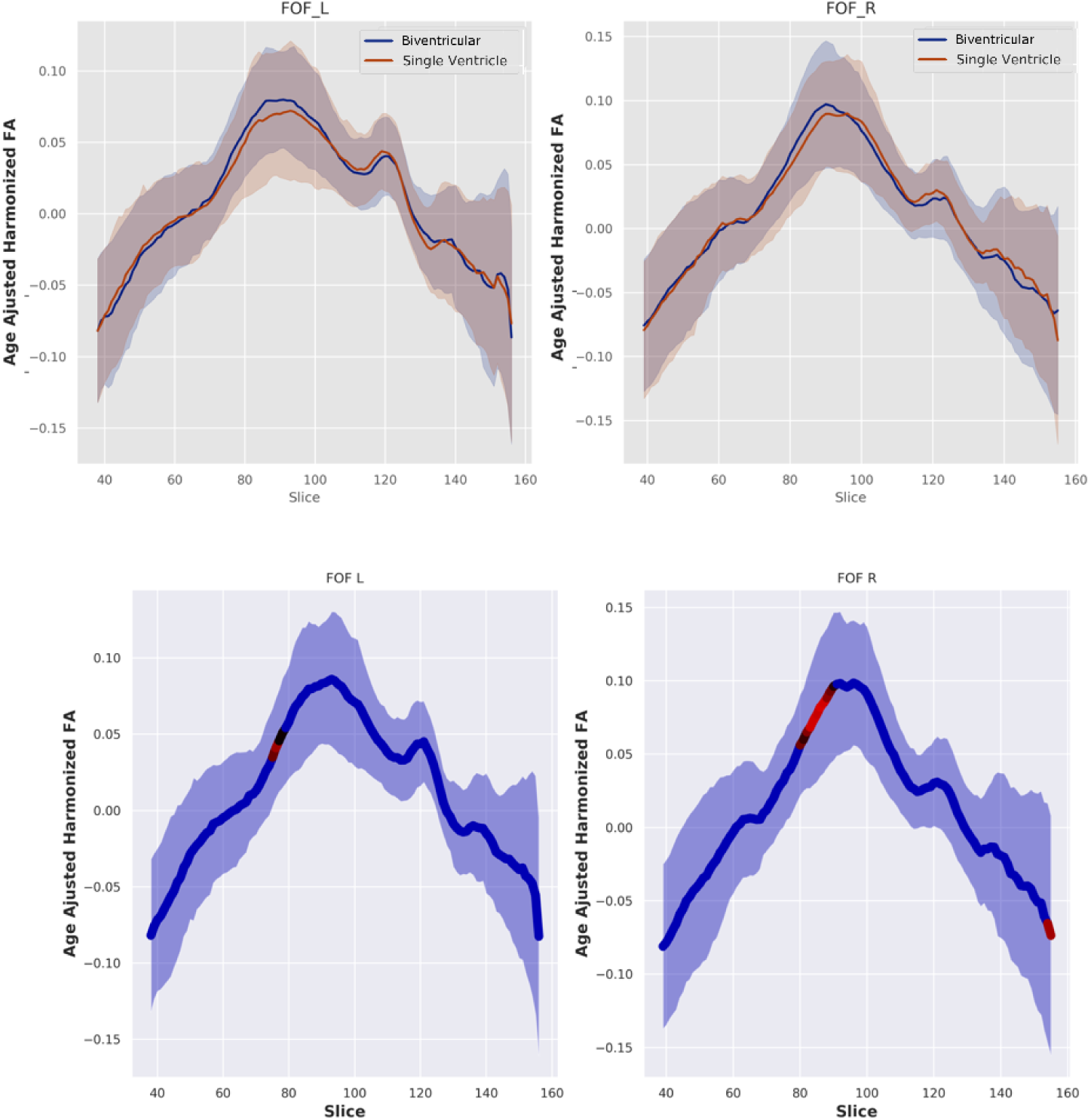
Post-operative along tract age-adjusted FA that was harmonized with ComBat in Fronto-Occipital Fasciculus (FOF). A) mean distribution between single vs. bi-ventricular repair groups. B) Statistically significant differences (p < 0.05) between single and biventricular repair projected onto mean along-tract FA in red.

**Figure 13.**
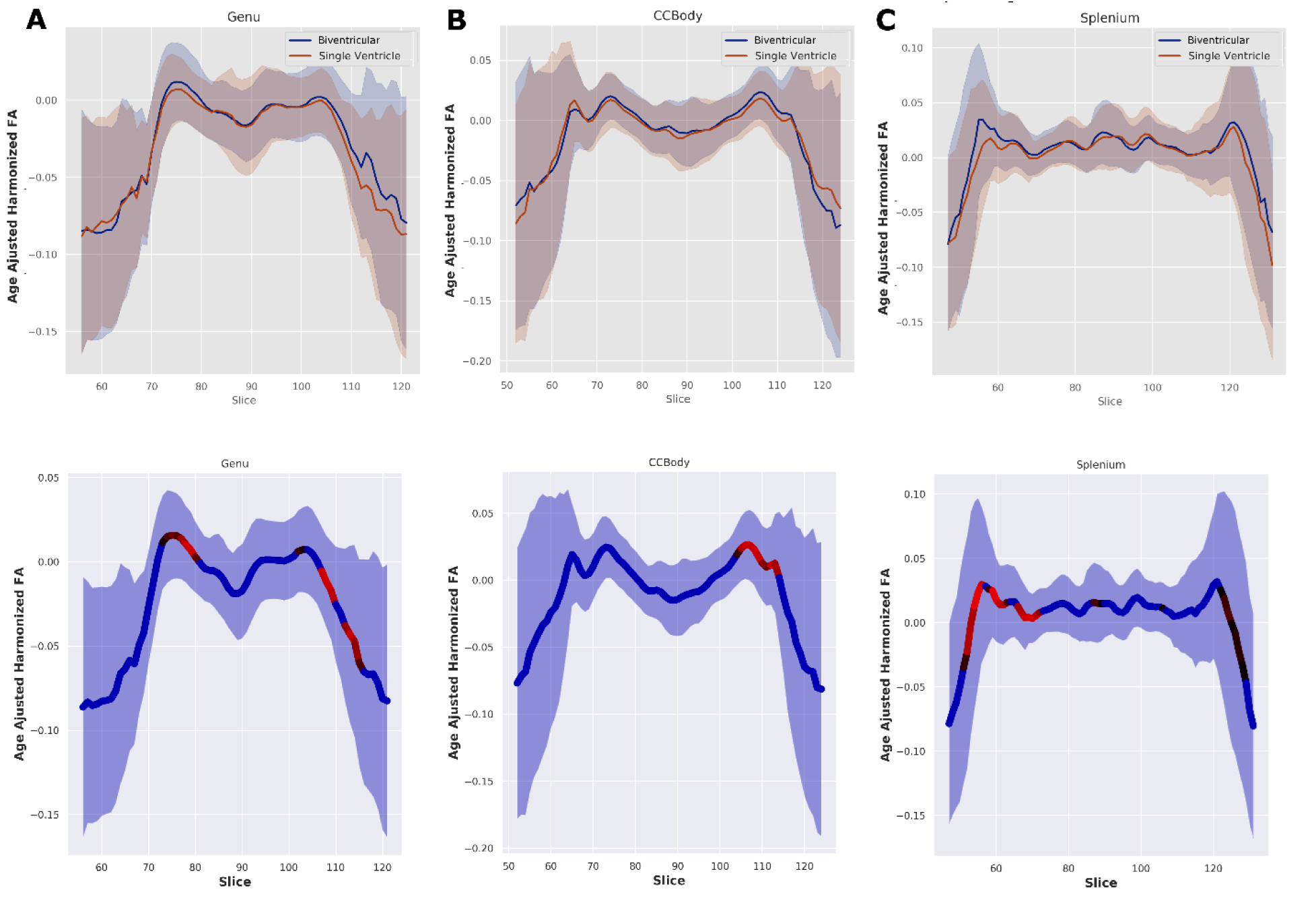
Post-operative along tract age-adjusted FA that was harmonized with ComBat in A) Corpus Callossum Genu, B) Corpus Callosum Body, and C) Corpus Callosum Splenium. Top row shows the mean distribution between single vs. bi-ventricular repair groups, bottom row shows the statistically significant differences (p < 0.05) between single and bi-ventricular repair projected onto mean along-tract FA in red.

### Availability and Computational Time

The full Dockerized pipeline is freely available at https://github.com/PIRCImagingTools/DINGO. On an Intel® Core™ i9-7900X CPU @ 3.30GHz with 64 GB of memory the automated aspects of pre-processing will run approximately 3-4 minutes per scan. Though each registration when determining the representative subject took no longer than 3-5 minutes, there are N x N registrations, so this can potentially be the most time consuming. Finally, the ANTS registrations take approximately 2 hours per scan with the following call, ANTS 3 --image-metric CC[template, native, 1, 5] --number-of-iterations 50×50×50×50 --output-naming ANTS_OUT_ --regularization Gauss[0.0,3.0] --transformation-model SyN[25,3,0.05] --use-Histogram-Matching 0.

## Discussion

We have presented a framework for the analysis of neonatal diffusion tractography imaging, with an emphasis on retrospective harmonization of multi-site diffusion tensor imaging [multi-vendor, different field strengths (1.5T and 3T) and different acquisitions] of a high-risk neonatal population (CHD). We chose to build our pipeline with explicit emphasis on the uncertainty of the estimated group parameters (diffusion metrics and spatial similarity) by having automated outlier detection and site- and group-wide harmonization prior to data analysis. This allows for the building of cleaner, more rigorous statistical models able to detect smaller effect sizes in heterogeneous populations. Additionally, to our knowledge, this is the first time ComBat (empirical Bayes harmonization) has been applied to neonatal tractography, and also to a high-risk clinical population. Using this technique, we show increased sensitivity to local changes in white matter tracts pre- and post-surgical intervention in a large, multi-site population of neonates born with CHD.

Currently, there is no standardized approach to neonatal diffusion analysis. Individual workflows are tailored to a particular protocol^50,51^ or dataset. Under appropriate conditions, protocol-specific pipelines offer the best approach for data extraction and quantitation. However, this approach is not scalable, or can introduce site-specific artifacts in heterogeneous data. In the setting where dealing with heterogeneous data is required, we need a framework that can scale to large data analysis while handling the intrinsic scanner- and site-specific variance. A popular alternative to manual delineation, and one of the most widely used group-level methods of DTI analysis, is Tract-Based Spatial Statistics.^41,52,53^ TBSS is an approach that transforms all subjects in the analysis into one common space, and projects each subject’s spatial diffusion metrics onto an atlas-derived tract “skeleton”. TBSS is effective at analyzing global differences between large groups, and is generally robust in large tracts with low spatial variance among subjects. In effect, TBSS looks only at the most prominent fibers within the group and restricts the generation of the FA skeleton based on overall FA values. This has two significant downsides, especially in neonatal populations: 1) the non-linear transformation into a common space may distort a subject’s anatomy, leading to erroneous adjacent tract information propagating onto a skeleton, and 2) the restriction of the skeleton generation based on FA leads to a sub-optimal skeleton in neonates. Modified variants of TBSS have successfully improved its performance in neonatal datasets, but they still lack the local specificity that anatomically informed fiber tracking methods provide; and are still prone to errors in the context of anatomically abnormal subjects.^53^

Other variants of group-level approaches are more generalized atlas space methods. Similar to TBSS, these approaches transform the subject’s DTI into atlas space prior to tractography.^54,55^ This allows for the generation of one set of ROI/As, and all fiber tracking is performed in this common space. This approach removes the time-consuming step of manually delineating the regions for each individual subject. Additionally, atlas-based methods inherently provide inter-subject correspondence derived from the non-linear transformation onto this common space. Performing tractography in atlas space has some disadvantages, however. The non-linear transformation can distort the native curvature of more complex tracts. We may lose informative features of the native structure of the tract. Additionally, atlas-based approaches tend to be highly sensitive to abnormal structures, which can result in the exclusion of subjects due to potentially informative pathology. Therefore, we believe native-space along-tract analysis to be an effective compromise, with the search space reduction benefit of anatomically-driven tract generation, while standardizing the approach by propagating masks from a template.

Using DINGO, the inclusion of subjects with similar structural abnormalities by grouping them into their own sub-cohort specific template is likely to improve tractography accuracy, retaining potentially informative pathology, reducing analysis bias, and retaining statistical power. We implement the outlier removal step at the end of the workflow as a way of empirically identifying and removing subjects from the cohort without user bias prior to analysis. Isolation Forests are data agnostic, and can identify outliers independent of anatomy, pathology, or visual differences. We believe this approach provides a way of utilizing a larger portion of abnormal populations in the analysis, with a worthwhile tradeoff of a small added computational time. Our implementation of IF detected a small subset of outliers (5 pre-op, 8 post-op). This small number was expected, as we chose a conservative approach for outlier detection as to not exclude too many subjects from the abnormal population, and a prior selection bias was already in effect due to pre-processing QA and the selection of subjects stable enough to undergo an MRI. Despite this low number of outliers, we noticed an increase (not statistically significant) in cross-site variance after harmonization when the outliers detected by this algorithm were included in the dataset.

Evaluating our pipeline on a heterogeneous dataset of neonates with CHD acquired at multiple institutions presents an ideal use case under controlled conditions – the images were acquired at multiple institutions, but under the same clinical trial surgical protocol^35,56^. Thus, it enables us to explore our null hypothesis that there should be no observed differences across similarly stratified subjects, while retaining the power to detect a significant effect size within the strata. Here we dichotomized the subjects as single vs. bi-ventricle CHD. Prior to harmonization, we detected larger cross-site effect sizes than the between-strata effects. After harmonization, we observed an amplified effect size when comparing single vs. bi-ventricle, with a decrease in the cross-site variance. We observed larger changes in white matter diffusivity between single vs. bi-ventricle after surgical intervention. This may reflect a direct biological signal, and it has been previously observed in similar populations that there is a compounding injury effect observed over time using structural imaging.^57^ However, we cannot rule out that we are more likely to detect subtle white matter changes at later time points, as increased diffusion signal in the developing brain allows us to tract both central and peripheral tracts with more fidelity. In this context, this work highlights the importance of data harmonization in multi-site studies. We are able to build more sensitive models where we can reduce the contribution of site and scanner variance.

This study has several limitations. Implementing a semi-automated pipeline inherently comes with trade-offs, compared to more manual approaches. First, automated tractography requires larger ROI/As to account for anatomical variance across the population, in order to more robustly delineate the tracts of interest. We carefully constructed our templated ROI/As to account for the potential for spurious tracts, adding additional regions of avoidance where necessary. Additionally, we refrain from “tract pruning” corrections, as these steps can often introduce bias. Our future work will include applying DINGO to independent neonatal datasets, which will allow us to use machine-learning to guide objective quality assurance and identify further technical outliers, including population-based FA cut-off, fiber bundle thresholding, and using gray and white matter masks to restrict tract pathways. We chose to perform our along-tract analysis by following the primary tract direction in an atlas space, rather than individual curvature-based approaches.^19^ This can lead to a loss of spatial information, particularly for high curvature tracts. However, in a heterogeneous neonatal population, following individual tract curvature can be noisy, as these curvature approaches can be unreliable due to lower structural resolution (SNR) and anatomical variance. Finally, ComBat harmonization, despite outperforming conventional methods, is limited in smaller datasets. By design, a different set of corrective values dependent on co-variates of interest needs to be generated with each statistical analysis. If the sample size is sufficiently large, this has low impact on data analysis. This is advantageous for large population studies, where we have enough confidence that the true population variance is modeled by the available data, and the Empirical Bayes method of estimating the priors directly from the data being analyzed is acceptable. Therefore, when we are restricted to retrospectively analyzing a multi-site dataset, without further access to data acquisition at each site, empirical Bayes outperforms classical statistical approaches, and improves statistical power.^2,48^

## Data Availability

Data is available upon request.

https://github.com/PIRCImagingTools/DINGO

## Supplemental Material

**Supplemental Figure 1.**
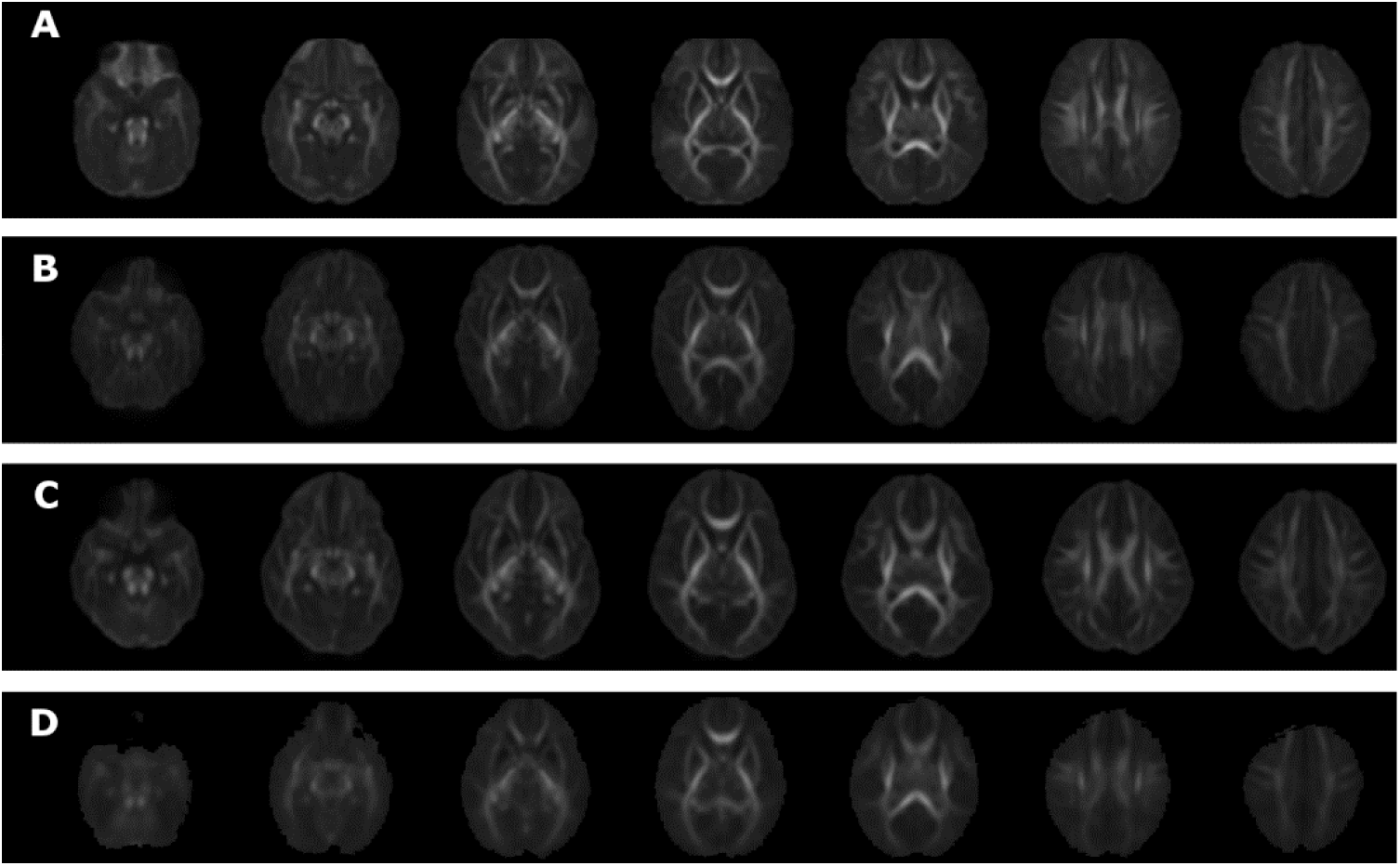
Site-specific mean FA in most representative space. A) CHLA, B) CHoP, C) CHP, D) TCH.

**Supplemental Figure 2.**
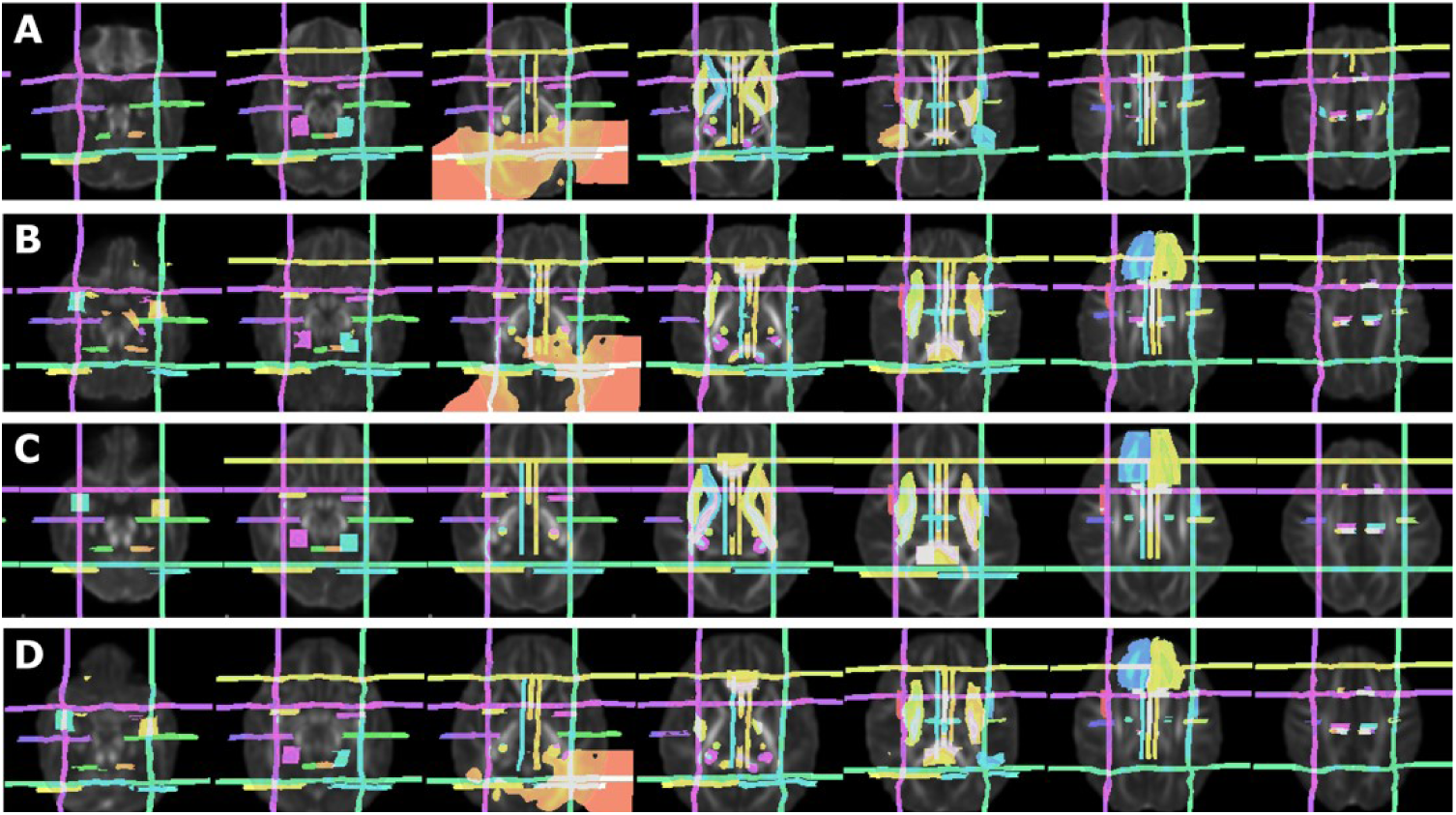
Cohort generated masks in template space. A) CHLA, B CHoP, C) CHP, D) TCH.

**Supplemental Figure 3.**
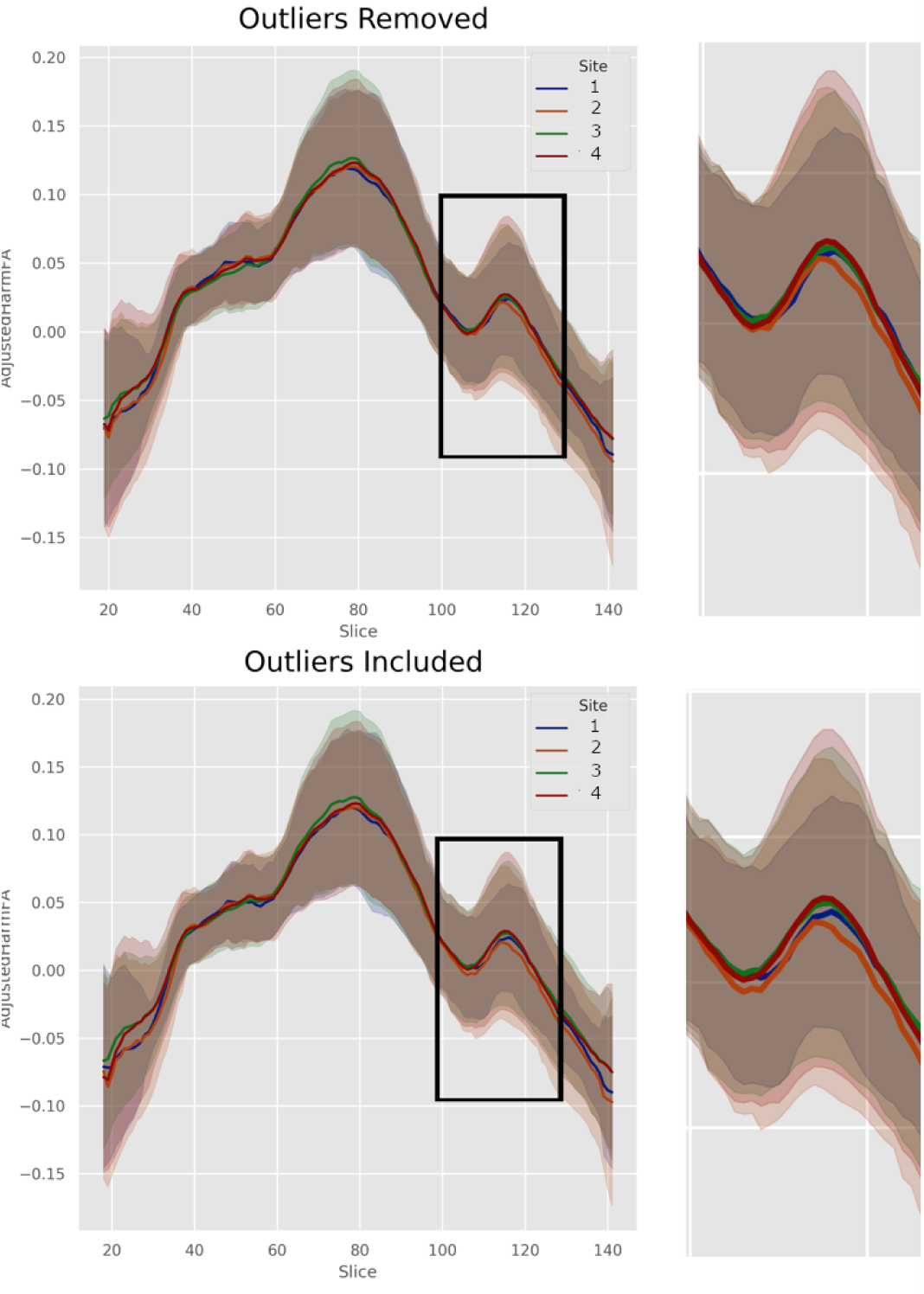
Comparison of the inclusion of 5 outliers identified by Isolation Forest algorithm in the final harmonization step in the left corticospinal tract at post-operative time point. Noticeable increase in cross-site variance can be observed in regions of lower signal and crossing fibers.

**Supplemental Table 1.**
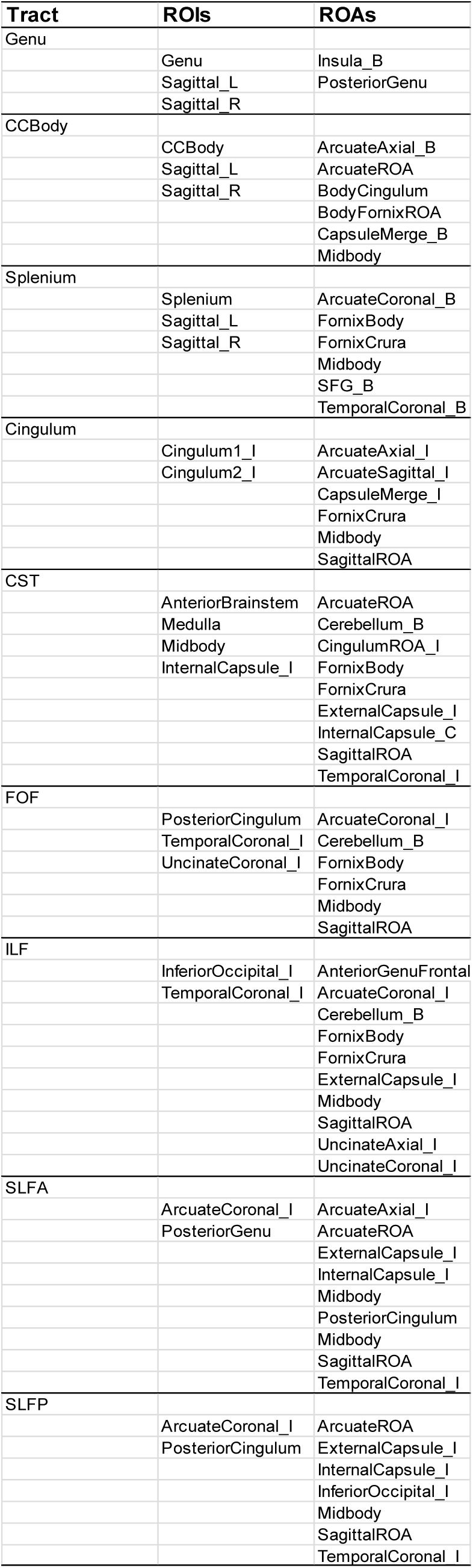
Mask Set description for each delineated tract.

